# Evaluation of Urine SARS-COV-2 RT-PCR as a predictor of Acute Kidney Injury and disease severity in critical COVID-19 patients

**DOI:** 10.1101/2021.01.13.21249576

**Authors:** Sérgio Pinto de Souza, Marcelo Augusto Duarte Silveira, Bruno Solano de Freitas Souza, Carolina Kymie Vasques Nonaka, Erica de Melo, Julia Cabral, Fernanda Coelho, Rogério da Hora Passos

## Abstract

The novel coronavirus disease (COVID-19) is an emerging infectious disease caused by SARS-CoV-2, which began as an outbreak in Wuhan, China and spread rapidly throughout the globe. Although the majority of infections are mild, severe and critical COVID-19 patients face deterioration of respiratory function, and may also present extrapulmonary manifestations, mostly affecting the kidney, digestive tract, heart and nervous system. Here, we prospectively evaluated the presence of SARS-CoV-2 genetic material by RT-PCR in urine samples obtained from critical care COVID-19 patients. In 51 patients included, we found higher serum creatinine levels, a longer hospital stay and a more frequent dialysis need in urine-positive patients. These findings could suggest that, in predisposed patients, a direct viral cytopathic effect may contribute to a more severe disease phenotype.

## Introduction

The novel coronavirus disease (COVID-19) is an emerging infectious disease caused by SARS-CoV-2, which began as an outbreak in Wuhan, China and spread rapidly throughout the globe. Although the majority of infections are mild, severe and critical COVID-19 patients face deterioration of respiratory function, and may also present extrapulmonary manifestations, mostly affecting the kidney, digestive tract, heart and nervous system^1^. Acute kidney injury (AKI) is one of the important complications of the COVID-19, with an overall incidence of 4.5%, but affecting up to 36% of patients with severe disease^2^. AKI is considered a marker of disease severity and a negative prognostic factor for survival^3^.

Extrapulmonary manifestations may be explained by direct infection of cells in target organs, as the kidney, since viral dissemination may occur through the bloodstream, leading to invasion of the organ and injury to the resident renal cells^4^. Other plausible mechanisms are inflammation-driven, since such patients face cytokine storms, with systemic inflammation-mediated injury, and are frequently dehydrated and exposed to secondary infections in the intensive care unit^5^. The direct viral cytopathic effect hypothesis is supported by previous reports of detection of viral genetic material in the blood and urine samples obtained from COVID-19 patients^6^, as well as by evidence from post-mortem studies that showed the presence of viral particles in the renal tissue^7^. Also, renal abnormalities as hematuria and proteinuria are commonly reported, and associated with higher mortality^8^. However, the clinical relevance of the coronavirus-induced cytopathic effect in the AKI of COVID-19 remains unclear.

Here, we prospectively evaluated the presence of SARS-CoV-2 genetic material by RT-PCR in urine samples obtained from COVID-19 patients admitted in the intensive care units in Hospital São Rafael in Salvador, Brazil. We intended to investigate a possible association between positive results of SARS-CoV-2 RT-PCR in urine samples and AKI onset, and abnormalities in urine sediments. Our hypothesis was that the virus would be detected more frequently in patients that develop AKI, supporting the role for a direct cytopathic effect in the renal parenchymal as the underlying mechanism of AKI in COVID-19 patients.

## Methods

### Ethics statement

The study was conducted following the principles of the Declaration of Helsinki and received prior approval by the Ethics Committee of São Rafael Hospital in Salvador, Brazil (CAAE 34428920.0.0000.0048). All participants gave informed consent.

### Subject selection

All adult patients with suspected COVID-19 pneumonia requiring oxygen supplementation admitted to the intensive care unit of São Rafael Hospital between July 10th and September 16th 2020 were screened for eligibility. Patients were included in the study if they received the diagnosis of COVID-19 pneumonia confirmed by nasopharyngeal RT-PCR and chest CT scan. Fifty-one subjects were included in the study and urine samples were collected in forty-nine patients. AKI was diagnosed by KDIGO criteria.

### Isolation of viral RNA by RT-PCR

SARS-CoV-2 RNA was detected in the nasopharyngeal swab samples using a quantitative real-time RT-PCR. Urine samples were stored at -80°C before nucleic acid extraction and real-time RT-PCR. Urine nucleic acid extraction was performed using NucleoSpin RNA for RNA purification with an elution volume of 50 µL (Macherey-Nagel, Düren, Germany). Real-time RT-PCR was performed using the Allplex™ 2019-nCoV Assay (Seegene, Seoul, Korea) that detects the following targets: RdRP gene and N gene specific for SARS-CoV-2, and E gene for all of Sarbecovirus, in a single tube. Cycle thresholds below 40.0 were considered positive by the 7500 real-time PCR System (Thermo Scientific, Waltham, MA, USA).

### Data analysis

Categorical variables were compared using fisher exact test. Continuous data were presented as median and interquartile range; the Mann-Whitney U test was used for comparisons. P values < 0.05 were considered significant. Data was analyzed with the PSPP® statistical package.

## Results

Prospective follow up of the participants revealed an AKI incidence of 50.9%. AKI onset occurred during the first days of admission in the ICU or at the study enrollment in most of them. RT-PCR analysis of urine samples revealed a positive result in six patients (12.2%) and no correlation was found between AKI and viral RNA detection in the urine by RT-PCR. Clinical and laboratorial data are shown in table 1. We found a statistically significant difference by comparing the frequency of comorbidities - hypertension and diabetes mellitus - in the group with positive RT-PCR result in urine samples compared to the group with negative results. Patients with positive SARS-CoV-2 RT-PCR results in urine samples also presented significantly higher creatinine levels at admission and presented the highest creatinine values during hospital stay (Table 1). Proteinuria was found to be more common in these patients (66.6% vs 46.6%), as well as the incidence of AKI (66.6% vs 48.9%). A second urine sample was collected in three patients, and was negative in all of them. Patients presenting positive RT-PCR result in urine samples also presented a longer hospital stay.

**Table 1.**
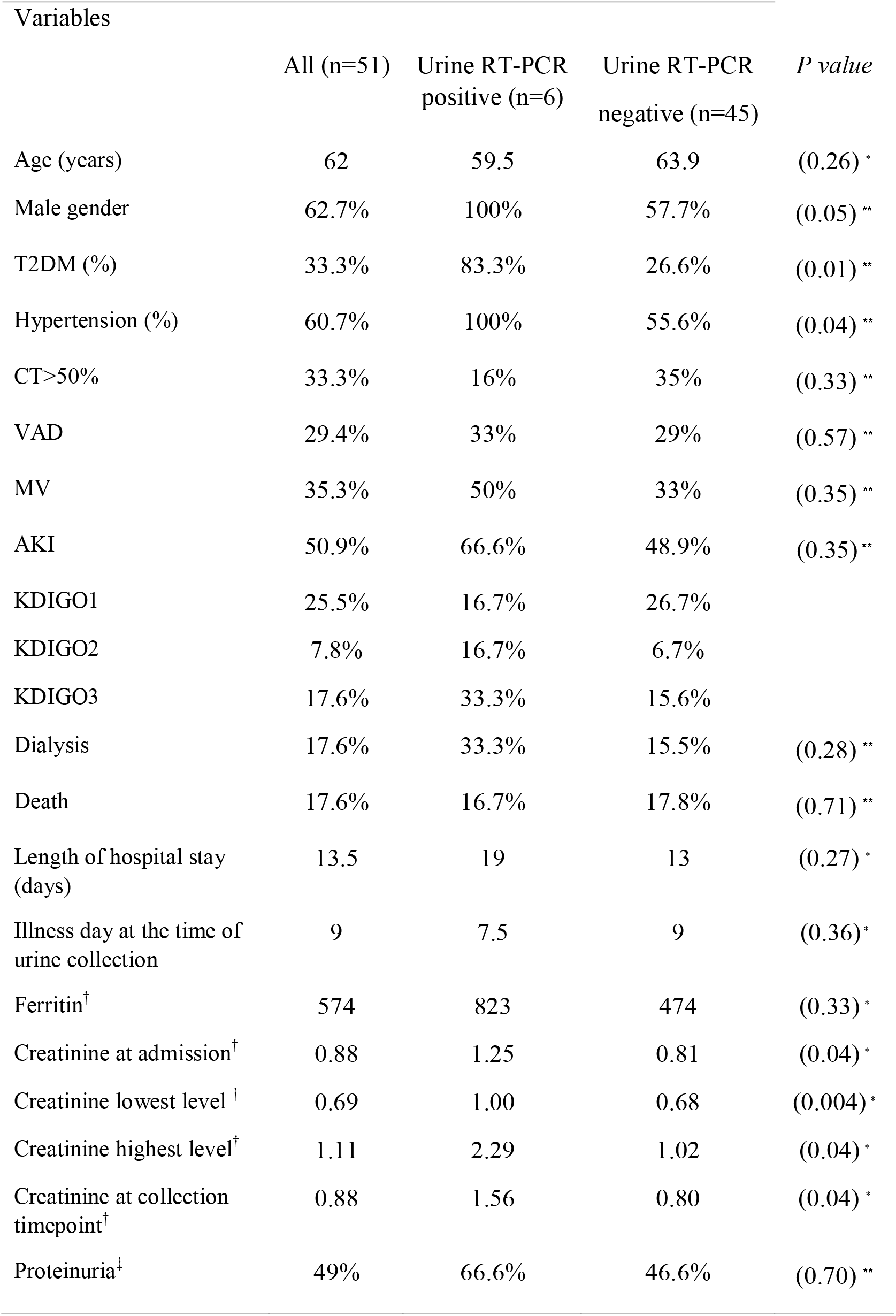
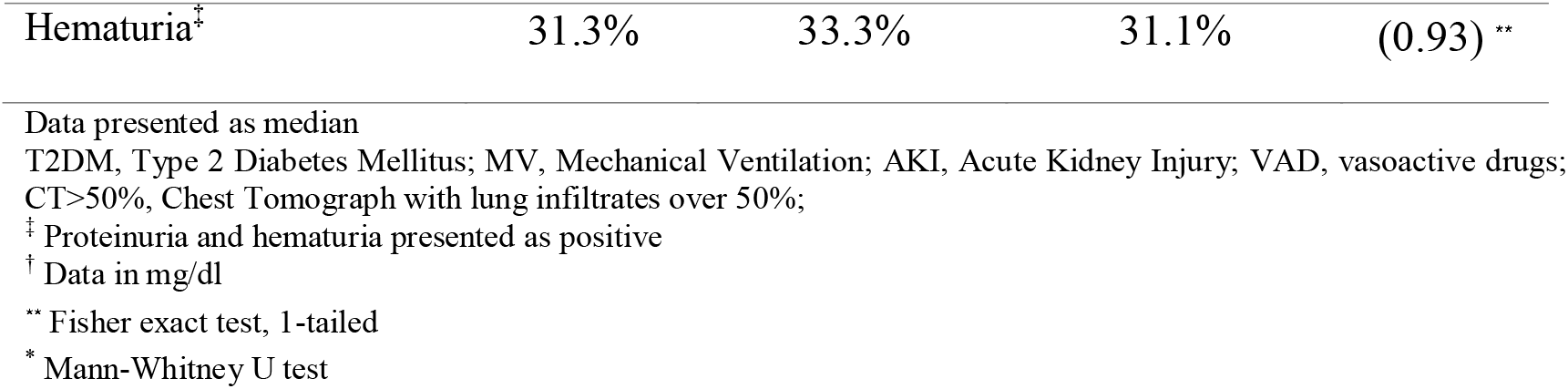
Clinical and laboratory variables of the patients with COVID-19 according to urine RT-PCR.

## Discussion

In this prospective cohort, we found that a positive RT-PCR in the urine was an infrequent finding. These patients were all male, had higher prevalence of diabetes and high blood pressure, that are well known risk factors for severe disease. They also had more urine proteinuria, higher serum creatinine levels and a higher prevalence of AKI, in accordance to previous reports^2^. Taken together, these findings may suggest a possible cytopathic viral effect influencing the severity of renal disease.

Also, the duration of symptoms was shorter in the group with detected SARS-CoV-2 in urine samples. To the best of our knowledge, another study reported a lower detection of the viral RNA in the urine, but the median days of symptoms before collection was fourteen days^9^. So, we speculate that viremia, as well as renal cells infection and associated viral shedding in the urine could be an early event.

In addition, our findings also point to multifactorial causes already described in the IRA related to COVID-19, such as exposure to nephrotoxins, volemic state, systemic inflammation and endothelial lesions (microthrombi formation)^10^. This hypothesis could explain the relatively low prevalence of proteinuria in the urine negative patients. Also, only one third of all patients presented hematuria; this finding, also, does not support a relevant role for direct viral-mediated cytopathic effects the pathophysiology of AKI in a high proportion of COVID-19 patients.

Our study has some limitations. It was a single center, observational study with a limited number of participants. We also did not successfully obtain follow-up urine specimens, as the majority of AKI patients already had AKI upon enrollment in the study.

In summary, we searched prospectively the urine of COVID-19 patients for the SARS-CoV-2 RNA, and found a relatively low incidence of this finding, which does not support a relevant role for cytopathic viral effects in the majority of AKI patients. However, the subgroup of patients who tested positive for SARS-CoV-2 in urine samples presented a higher frequency of diabetes and hypertension. In this subgroup, the association of higher serum creatinine levels, a longer hospital stay and a more frequent dialysis need were found. These findings, although preliminary, lead to the speculation that, in predisposed patients, a direct viral cytopathic effect could contribute to a more severe phenotype.

### Disclosure

All the authors declare no competing interests.

## Data Availability

All data referring to the manuscript are available.

